# Initial Analysis of Viral Dynamics and Circulating Viral Variants During the mRNA-1273 Phase 3 COVE Trial

**DOI:** 10.1101/2021.09.28.21264252

**Authors:** Rolando Pajon, Yamuna D. Paila, Bethany Girard, Groves Dixon, Katherine Kacena, Lindsey R. Baden, Hana M. El Sahly, Brandon Essink, Kathleen M Mullane, Ian Frank, Douglas Denhan, Edward Kerwin, Xiaoping Zhao, Baoyu Ding, Weiping Deng, Joanne E Tomassini, Honghong Zhou, Brett Leav, Florian Schödel, on behalf of the mRNA-1273 COVE Trial Team

## Abstract

This analysis assessed the impact of mRNA-1273 vaccination on the viral dynamics of severe acute respiratory syndrome coronavirus-2 (SARS-CoV-2) infection in the ongoing Coronavirus Efficacy (COVE) trial. mRNA-1273 vaccination significantly reduced SARS-CoV-2 viral copy number (95% confidence interval [CI]) by 100-fold on the day of diagnosis (4.1 [3.4-4.8] versus placebo (6.2 [6.0-6.4] log10 copies/ml). Median times to undetectable viral copies were 4 days for mRNA-1273 and 7 for placebo. Vaccination also reduced the burden of disease and infection scores. Vaccine efficacies (95% CI) during the trial against SARS-CoV-2 variants circulating in the US were 82.4% (40.4%-94.8%) for Epsilon and Gamma, and 81.2% (36.1%-94.5%) for the Epsilon variants. The detection of other respiratory viruses during the trial was similar between groups. In those who became SARS-CoV-2 infected, the reduction of viral load after mRNA-1273 vaccination is potentially correlated to the risk of transmission, which has not been assessed in this study.

## Introduction

The mRNA-1273 vaccine, a lipid nanoparticle-encapsulated messenger RNA vaccine encoding a prefusion-stabilized spike (S) protein of the prototype Wuhan-Hu-1 virus isolate, demonstrated high efficacy in preventing symptomatic and asymptomatic severe acute respiratory syndrome (SARS-CoV-2) infections at the primary analysis (December 2020) of the ongoing Coronavirus Efficacy (COVE) phase 3 trial.^1,2^ Following Emergency Use Authorization [EUA] issuance of mRNA-1273 in the United States (US), the trial protocol was amended from the observer-blind part of the study to an ongoing open-label part. A recent update of the results reported after completion of the blinded portion of the study (March 2021) with 5.3 months of follow-up time, showed a vaccine efficacy (VE) rate of 93.2% and safety profile similar to those observed at the primary efficacy analysis after 64 days.^2^

While SARS-CoV-2 vaccines are highly effective against symptomatic and asymptomatic infections, less is known about the effect of vaccination on viral load in the upper airways of persons with breakthrough infection, and the duration and magnitude of viral shedding. Viral load in the upper airways is an important marker of SARS-CoV-2 infection, and is related to the severity and symptoms of coronavirus disease 2019 (Covid-19), clinical outcomes and mortality, as well as SARS-CoV-2 transmission.^3-10^ Recent prospective and retrospective studies have reported reductions in viral load and duration of viral shedding following Covid-19 vaccination^11,12^

The emergence of several SARS-CoV-2 variants with mutations in the S protein genes and in other regions of the genome with decreased susceptibility to neutralization by vaccine-induced antibodies has raised the possibility for increased transmission, breakthrough infections and waning efficacy with current vaccines.^13-22^ The variants include those previously considered to be variants of concern (VOC; Alpha [B.1.1.7], Beta [B.1.351], and Gamma [P.1]), and variants of interest (VOI; Eta [B.1.525], Iota [B.1.526], Kappa [B.1.617.1] and Epsilon [B.1.427, B.1.429]) that continue as variants being monitored.^23^ From initiation of the COVE trial (July 2020) to completion of the blinded portion of the study (data cutoff-date of March 26, 2021), the proportions of circulating VOC and VOI were very low in the US and were predominantly of the Alpha, Beta, Epsilon and Iota lineages. In recent months, the highly transmissible Delta (B1.617.2) VOC, has become the predominant cause of SARS-CoV-2 infections in the United States.^23-26^ Co-infection of SARS-CoV-2 with respiratory pathogens also occurs, and can complicate the evaluation of SARS-CoV-2 infection rates as well as patient management due to the presence of non-specific symptoms unrelated to Covid-19 illness.^27-29^

The aim of this study was to assess the effect of mRNA-1273 vaccination on viral copy and viral shedding, as well as the burden of disease in participants with Covid-19 by study group in the COVE ongoing trial. We also assessed the prevalence of SARS-CoV-2 viral variants and co-infecting respiratory pathogens, and the effect of vaccination on viral variants detected during the placebo-controlled phase of the study.^1^

## Methods

### Trial Design

This is an analysis of the previously reported COVE study, a phase 3 randomized, observer-blinded, placebo-controlled trial that enrolled adults in medically-stable condition at 99 US sites (clintrials.gov NCT04470427).^1,2^ Eligible participants included adults 18 years or older with no known history of SARS-CoV-2 infection, whose circumstances put them at appreciable risk for SARS-CoV-2 infection and/or high risk of severe Covid-19. Participants were randomized 1:1 to receive mRNA-1273 vaccine (100 µg) or placebo and stratified by age and Covid-19 complications risk criteria (≥18 and <65 years and not at risk, ≥18 and <65 years and at risk, and ≥65 years). Following issuance of the Emergency Use Authorization of mRNA-1273, the protocol was amended as a two-part phase 3 study (A and B; supplementary protocol). Part A was the observer blinded to-treatment phase which concluded when participants unblinded and consideration given those on placebo to receive mRNA-1273. Part B is the currently ongoing open-label phase. The blinded part of the trial was completed.^2^ Participants will continue to be followed-up for up to two years as originally planned.

The COVE trial is conducted in accordance with the International Council for Technical Requirements for Registration of Pharmaceuticals for Human Use, Good Clinical Practice Guidance, and applicable government regulations. The central Institutional Review Board/Ethics Committee, Advarra, Inc., 6100 Merriweather Drive, Columbia, MD 21044 approved the protocol and consent forms. All participants provided written informed consent.

The trial design, efficacy assessments and study treatment have been previously described and are provided in the supplementary protocol online.^1,2^ Briefly, the primary endpoint of the study was the vaccine efficacy of mRNA-1273 at preventing a first occurrence of symptomatic Covid-19 with onset ≥14 days post-second injection, Covid-19 cases defined as having ≥2 of systemic symptoms (fever ≥ 38ºC, chills, myalgia, headache, sore throat, new olfactory and taste disorder[s]), or experienced ≥1 respiratory sign or symptom (cough, shortness of breath, or clinical or radiological evidence of pneumonia), and confirmed by positive RT-PCR for SARS-CoV-2 using NP swab, nasal or saliva sample. The secondary endpoint, severe Covid-19, was defined as confirmed Covid-19 per the primary endpoint case definition, plus one of the clinical signs indicative of severe systemic illness (respiratory rate ≥30 per minute, heart rate ≥125 beats per minute, SpO2 ≤93% on room air at sea level or PaO2/FIO2 <300 mm Hg, OR respiratory failure or Acute Respiratory Distress Syndrome [ARDS, defined as needing high-flow oxygen, non-invasive or mechanical ventilation, or ECMO], evidence of shock [systolic blood pressure <90 mmHg, diastolic BP < 60 mmHg or requiring vasopressors], OR significant acute renal, hepatic or neurologic dysfunction, OR admission to an intensive care unit or death). Safety included evaluation of solicited local and systemic adverse events with onset during the 7 days following each injection to resolution, unsolicited adverse events during 28 days following each injection, adverse events leading to discontinuation from dosing and/or study participation, medically-attended and serious adverse events throughout the study, and severity graded as described in the protocol.

In this analysis, viral variants were sequenced and viral copy number and shedding were assessed in 799 adjudicated Covid-19 cases in the per-protocol set from the blinded portion of the study (data cut-off March 26th, 2021). Following amendment (December 23rd, 2020) of the COVE phase 3 study, the open-label phase of the trial was initiated and participants in the placebo arm started to receive the mRNA-1273 vaccine. Sequence data for viral variants and also assessment of the presence of other respiratory pathogens extended into the time-frame of the open-label part of the study.

### Study procedures

#### Viral load assessment

##### Study design and population

The study population for analysis of viral load were those participants in the per-protocol population who had binding antibody (Elecsys; nucleocapsid protein [NP]) negative baseline values (baseline SARS-CoV-2 negative), and also had follow-up Elecsys-negative values at days 29 and 57 and RT-PCR-negative results at baseline, day 29, and at day 57 (only Elecsys). The analysis period was limited to the blinded portion of the study and the data cut-off date was 26-March-2021 (or earlier unblinding).

##### Statistical Analysis

For the cohort of Covid-19 adjudicated subjects, the day 1 illness nasopharyngeal swab and the days 3, 5, 6, 9, 14, 21, and 28 of illness saliva specimens were matched to assess the qualitative and quantitative results for each. SARS-CoV-2 RT-PCR was performed as described below (Eurofins Viracor, Kansas City, MO). Conversion from cycle-threshold (Ct) time to viral copies for the quantitative RT-PCR were Log10 viral copies/mL=Ct-40.9578)/-3.3385 for swabs (day1) and Log10 viral copies/mL=CT-41.0349)/-3.3346) for saliva (days 3, 5, 7, 9, 14, 21 and 28). If the qualitative result was negative, the log10viral copies was assumed to be 0. A mixed model repeated measures (MMRM) analysis was performed comparing absolute and change from baseline log10 viral load between vaccinated and placebo participants from the nasopharyngeal swab day 1 of illness through saliva sample days 3, 5, 7, 9, 14, 21, and 28 of illness. Only participants with a quantitative result in the day 1 illness nasopharyngeal swab were included in the MMRM modeling. Any MMRM result estimate under 0 copies, was truncated at 0. There was no imputation for missing data.

#### Vaccine Efficacy Against Burden of Disease

An exploratory analysis of burden of disease (BOD) due to Covid-19 was performed in the PP set based on adjudicated cases in participants who were SARS-CoV-2-naîve by serology and PCR at randomization and had available post-baseline data. A BOD score was based on post-SARS-CoV-2 infection follow-up and defined to reflect the severity of symptoms (0 [without COVID-19] 1 [Covid-19 without hospitalization], 2 [Covid-19 with hospitalization], 3 [death]). A summary of BOD score and the number and percentage of participants with each level of BOD score was provided by treatment group. To assess disease burden in participants with Covid-19, a summary of BOD was provided in participants with Covid-19, (ie, participants with BOD score of zero were excluded from the analysis) and for assessment of the impact of baseline risk of severe disease on the vaccine effect regarding disease severity, a summary of BOD was provided by randomization strata (ie, ≥65 years; <65 years at risk; and <65 years not at risk). A proportional means model, including treatment group as fixed effect and stratified with stratification factor at randomization, was used to assess the vaccine effect on BOD. The VE for the BOD score was estimated as 1–the ratio of means as estimated by the proportional means model and reported with 95% CIs.

#### Vaccine Efficacy Against Burden of Infection

An exploratory analysis of burden of infection (BOI) was performed in the PP set based on asymptomatic infections and adjudicated Covid-19 cases in participants who were SARS-CoV-2 infection negative at baseline and had available post-baseline data. As with the BOD, a BOI score was defined to assess the severity of symptoms (0 [No infection], 1/2 [Asymptomatic infection], 1 [Covid-19 without hospitalization], 2 [Covid-19 with hospitalization, 3 [death]). A proportional means model, including treatment group as fixed effect and stratified with stratification factor at randomization, was used to assess the vaccine effect on BOI. The VE for BOI was estimated as 1 – the ratio of mean BOI scores and reported with 95% CIs.

#### Sequencing Methodology and Sequence Data Analyses

Sequencing of the SARS-CoV-2 spike gene was attempted from all available SARS-CoV-2 RT-PCR positive nasopharyngeal samples (n=832) collected between July 2020 through May 2021 from participants in the blinded portion of the COVE trial. This corresponded to 791 participants, (n=720 placebo and n=71 mRNA-1273). For 41 participants, more than one sample was sequenced within the illness period. The analysis of these cases is ongoing and will be included in additional analyses/reporting at a later date. Sequencing data was generated using three different approaches in two different laboratories.

##### Spike-Gene Sequencing Assay from Eurofins Viracor (Kansas City. MO)

Viral RNA from nasal swabs was extracted using the NucliSENS® easyMag® extraction kit. Extracted RNA was used as template in a Qiagen One-Step Reverse Transcription-PCR (RT-PCR) reaction for cDNA synthesis using SARS-CoV-2 S gene Conventional RT-PCR primer mixes. The Agilent 2200 or 4200 TapeStation in conjunction with D5000 ScreenTapes, D5000 reagents, and the TapeStation Analysis software was used to assess post-amplified and purified PCR reactions for the presence, size, and concentration of any products generated. Library preparation was performed using Illumina Nextera XT Library Prep Kit. The Agilent 2200 or 4200 TapeStation in conjunction with D5000 ScreenTapes, D5000 reagents and the TapeStation Analysis software was used to assess purified libraries for presence, average fragment size, and concentration of the fragment distributions generated. Analysis of next generation sequencing data for the SARS-CoV-2 S gene NGS assay was done using Qiagen CLC Genomics Workbench v20.0.1 using NC_045512.2 as the reference strain. The custom workflow in CLC Genomics Workbench processed the sequencing data as follows: paired fastq files were imported, primer sequences were trimmed from 5’-ends of reads, reads were mapped to the full SARS-CoV-2 reference genome (NC_045512.2), single nucleotide and insertion/deletion variants relative to reference were called and annotated, and a consensus sequence of the spike gene (bases 21615 to 25436) was generated. The analysis workflow reported annotated variant tables, spike gene coverage tables, and spike-gene consensus sequences.

Upon TapeStation D5000 assessment and subsequent analysis of data using the TapeStation Analysis software, if the viral load was insufficient to obtain a correct band for the SARS-CoV-2 S gene targets (S1 (1026bp), S2 (893bp), S3 (1178), and S4 (1264bp), these results were considered negative. Positive results for the RT-PCR reactions were identified by 1) the presence of a band at the appropriate size for the SARS-CoV-2 S gene PCR products (S1 (1026bp), S2 (893bp), S3 (1178), and S4 (1264bp) relative to the D5000 ladder and 2) a peak table reporting a concentration for the specific bands for the sample. SARS-CoV-2 S gene NGS runs using MiSeq v2 chemistry reagents running paired-end 2 × 151 reads must exhibit the following criteria: Cluster densities approximating 600 - 1,200K/mm2; >80% of bases called exhibit Q-scores ≥30. Individual library sequence quality metrics were assessed by referencing the sequencing quality reports generated by analysis through the Qiagen CLC workbench program. For SARS-CoV-2 S gene NGS runs, up to 24 libraries can be sequenced on a single flow cell and the number of reads displayed in the trim summary section of the trim report should be ≥50,000 reads for each amplicon (prior to the reads being trimmed). Ninety-five percent of nucleotide positions between 21615-25436 should have a coverage of 100X. SARS-CoV-2 whole virus was used as a positive control. The LOD with all replicates for all 4 (S1-S4) amplicons was 6,667 copies/mL.

##### Spike-Gene Sequencing Assay from Monogram Biosciences (LabCorp, South San Francisco, CA)

Viral RNA from nasal swabs stored in UTM is extracted using the Kingfisher Flex platform. Following extraction, nested RT-PCR reactions are performed to amplify the entire SARS-COV-2 Spike (S) coding region. The resulting amplicon is purified and normalized before undergoing NGS library preparation (Kapa). Resulting barcoded libraries are normalized, pooled, and undergo 2×150 bp paired end sequencing on the Illumina MiSeq platform. FASTQ files are analyzed using a semi-automated bioinformatics pipeline. Briefly, reads are quality trimmed and overlapping paired reads are joined. Reads are aligned in a codon aware manner that maintains reading frame through the S gene. Codon and amino acid variants are determined and reported along with consensus sequences. Results are reviewed to ensure appropriate coverage levels and quality metrics were obtained for each run and each individual sample. A sensitivity to amplification of ∼1,000 copies/mL and a minor variant detection threshold of 3% was validated.

##### Whole Genome Sequencing Assay (WGS) from Eurofins Viracor (Kansas City, MO)

Viral RNA from nasal swabs is extracted using the Kingfisher Flex platform and GSD NovaPrime® RNA extraction kit. Extracted RNA is used as template in a one-step RT-PCR for cDNA synthesis. Each cDNA is subjected to amplification using ARTIC SARS-CoV-2 Primer Pools. These primer pools were designed to amplify approximately 90 amplicons each with each amplicon averaging ∼400bp. Mapping these amplicons to a reference sequence illustrates the ‘tiled’ approach used for primer design resulting in coverage of the entire SARS-CoV-2 genome. Purification of the ARTIC PCR reactions was performed manually with Beckman-Coulter SPRIselect magnetic beads. The concentration of amplified amplicons in each sample was quantified using the Qubit FLEX fluorometer. Preparation of libraries was performed using the NEBNext Ultra II FS library prep kit in conjunction with the BRAVO liquid handling platform. Automated purification of the library reactions was performed using the Agilent BRAVO liquid handler and Beckman-Coulter SPRIselect magnetic beads. The fragment size distribution of the final pooled library was confirmed using the Agilent TapeStation 4200 or ThermoFisher BioAnalyzer 2100 DNA fragment analyzer prior to preparation for sequencing. Pooled libraries were denatured and sequenced on the NextSEQ 500 or 550 instrument using a NextSEQ Mid Output 500/550 flow cell and reagents running a 2×150 cycle paired-end sequencing protocol. A Twist SARS-CoV2 RNA positive control was processed in parallel with each verification run for positive control of the RT-PCR, ARTIC PCR amplification, and library preparation. The LoD for SARS-CoV2 WGS was determined to be 100 copies/ARTIC PCR assay reaction.

#### Variant Data Analyses

SARS-CoV2 variants in the study population were assessed based on amino acid mutations in the spike protein relative to the reference strain (spike mutations). For each of the three sequencing datasets, (spike-gene sequencing from Eurofins Viracor; spike-gene sequencing from Monogram Biosciences (LabCorp, South San Francisco, CA), and whole genome sequencing from Eurofins Viracor), spike protein amino acid mutations were acquired directly from the sequencing service provider. For each specimen, a single spike haplotype was designated as the ordered set of spike mutations. The same analysis was performed for the global SARS-CoV-2 genomic database available from the Global Initiative on Sharing All Influenza Data (GISAID).^25,30^ Pango Lineages^31 30^ were inferred for the clinical specimens in two ways. First, they were inferred from the lineage annotations of matching spike haplotypes included in the GISAID database. In cases when a single spike haplotype was associated with more than one Pango Lineage in GISAID, the dominant lineage was used. In addition, Pango Lineages were inferred based on the presence of core backbone mutations from CDC variants in a specimen’s spike haplotype.^23^ To assess the relative prevalence of select lineages, the full clinical dataset and full GISAID dataset were subset to include only records annotated with the selected lineages. Prevalence for each selected lineages was then computed as the percentage of total records for a given month within each data subset.

##### Variant-specific Vaccine Efficacy of mRNA-1273 - Statistical Analysis Method

For specific viral variants with a sufficient number of variant cases during the blinded phase of the study, the competing risk method was used to estimate the vaccine efficacy of mRNA-1273, specifically, Fine and Gray’s (FG) sub-distribution hazard model was used. COVID-19 cases of specific variant were considered as cases, and COVID-19 cases of variants other than that of interest were considered as competing events in this analysis.

##### SARS-CoV-2 RT-PCR Test (Eurofins Viracor, Kansas City, MO)

SARS-CoV-2 specific real-time reverse transcription polymerase chain reaction (RT-PCR) assay was used to detect SARS-CoV-2 RNA in upper respiratory (nasal/nasopharyngeal wash and swab) and bronchoalveolar lavage (BAL) samples. This assay, performed by Eurofins Viracor Laboratories, Kansas City, MO provides qualitative detection of RNA from SARS-CoV-2 virus in specimens collected from individuals meeting SARS-CoV-2 virus clinical criteria (https://www.fda.gov/media/136740/download). Briefly, the SARS-CoV-2 RT-qPCR assay was performed as a multiplex reaction with the MS2 internal control assay. Oligonucleotide primers and TaqMan probes were used for the detection of two regions of the viral nucleocapsid (N) protein gene region of SARS-CoV-2 and an internal extraction and amplification control target (the RNA bacteriophage MS2) was used. The limit of detection (LOD) for this assay is determined to be 73 copies/mL for BAL, nasal wash and nasopharyngeal swab with a cycle threshold (Ct) of 38 being the cutoff for positive result. The SARS-CoV-2 RT-qPCR was also used for quantitative detection of viral RNA in both nasopharyngeal swabs and in saliva samples. The LOD is 299 copies/ml and lower level of quantitation (LLOQ) is 714 copies/ml.

#### Detection of Respiratory Pathogens

Participants with symptoms of a respiratory illness were advised to contact the clinical site within 72 hours of onset. Such visit defined the day 1 illness date and triggered SARS-CoV-2 molecular testing via a nasopharyngeal swab that included testing for additional respiratory pathogens (Biofire RP2) and a 28-day follow-up period with periodic sampling (SARS-CoV-2 RT-PCR) also at days 3, 5, 7, 9, 14, 21 and 28 using saliva samples.

##### BioFire Respiratory Panel (RP2) (Eurofins Viracor, Kansas City, MO)

Respiratory pathogens were detected using the BioFire Respiratory Panel RP, a diagnostic multiplexed nucleic acid test intended for the simultaneous qualitative detection and differentiation of nucleic acids from 20 viral and bacterial respiratory organisms. The disposable closed system pouch was run on the Filmarray® Torch system which lyses samples, extracts, and purifies all nucleic acids, and performs nested multiplex PCR. Endpoint melting curve data was used to detect target-specific amplicons and analyze data to generate a result for each analyte. Nasopharyngeal swabs were used as qualitative diagnostic assays for the detection of 20 different viruses and bacteria associated with respiratory tract infection. Assay results are provided as negative or positive for each pathogen in the BioFire Respiratory Panel RP.

## Results

### Effect of mRNA-1273 Vaccination on SARS-CoV-2 Viral Load and Detectability of Infection

Viral load was assessed in the cohort of participants with adjudicated Covid-19 cases (at an illness visit) in the per-protocol (PP) population during the blinded and placebo-controlled phase of the COVE study.^1,2^ The analysis population comprised adjudicated Covid-19 cases in the PP Set with no evidence of infection through day 57, (i.e. those with negative binding antibody [bAb] against nucleocapsid protein [NP, ROCHE Elecsys]) and RT-PCR at baseline and day 29, and negative bAb against NP at day 57. There were a total of 799 adjudicated cases starting 14 days after dose 2 with 744 in the placebo and 55 in the mRNA-1273 groups (Figure 1). The differential in cases between groups is due to the demonstrated >93% efficacy of this vaccine.^1,2^ Of the 799 adjudicated cases, 701 (48 in the mRNA-1273 and 653 in the placebo groups) had no evidence of infection through day 57 and were included in the analysis of viral load. Baseline demographics and characteristics were generally balanced for placebo and mRNA-1273 groups (Table S1). The mean age was 49 years (range 18-87 years) and 50% were female. In the mRNA-1273 group, more participants had chronic lung disease, significant cardiac disease, severe obesity and liver disease, lower incidences of diabetes and HIV, and a higher mean of body mass index than the placebo group.

**Figure 1.**
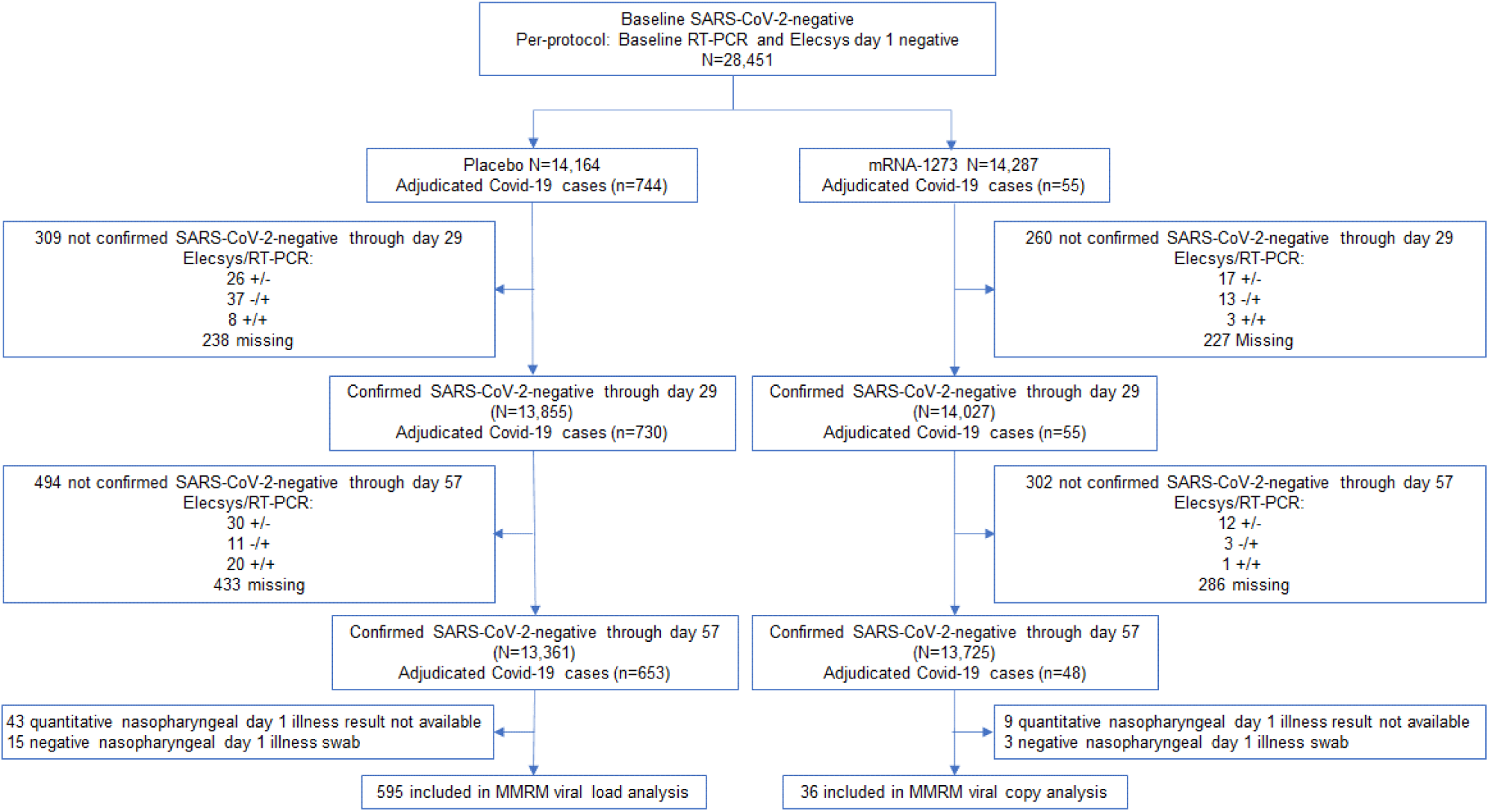
Viral copy and shedding analysis population. Included in the analysis were participants in the per-protocol population who were SARS-CoV-2 negative by both binding antibody against nucleocapsid (ROCHE Elecsys) and RT-PCR at baseline and day 29, and negative for binding antibody against nucleocapsid (ROCHE Elecsys) at day 57.^1^ The analysis was limited to adjudicated Covid-19 cases in the blinded portion of the study, i.e. earlier of unblinding or data cutoff date of March 26, 2021 based on a database lock which occurred on May 4, 2021.^1^

Viral load was assessed on the basis of SARS-CoV-2 2 RT-PCR Ct values converted to viral genome copy number. A MMRM analysis was performed comparing absolute and change from baseline in log10 viral load between vaccinated and placebo participants from nasopharyngeal swabs at day 1 of illness and in saliva samples through days 3, 5, 7, 9, 14, 21, and 28 of illness. Only participants with a quantitative result available at the day 1 illness nasopharyngeal swab were included in the MMRM analysis. A total of 36 participants in the mRNA-1273 and 595 in the placebo groups were included in the MMRM analysis of viral copy number. On illness visit day 1, the median number of viral copies/ml (log 10) were 6.7 for the placebo group and 3.4 for the mRNA-1273 group (Table S2). The MRMM analysis showed that from day 1 of illness through day 9, the number of viral copies detected in the placebo arm were significantly higher (p<0.001) than those in the mRNA-1273 arm (Figure 2A and Table S3). The difference (95% confidence interval) between the mRNA-1273 and placebo arms showed over a 100-fold reduction in viral copies/ml (log10) at day 1 (4.1 [3.4-4.8] and 6.2 [6.0-6.4]), and a 10-fold reduction at day 9 (0.06 [0-0.64] and 1.1 [0.9-1.2) in the mRNA-1273 group compared with the placebo group, respectively (Figure 2B and Table S4). Similarly, in age cohorts of participants 18 to <65 and ≥65 years of age, a 150-fold reduction in viral copy number at day 1 illness was observed for mRNA versus placebo (Figure S2). The median times to undetectable viral copies (lower limit of quantitation <2.85 log10 viral copies/ml) were 4 days for mRNA-1273 and 7 days for placebo (Figure S3).

**Figure 2.**
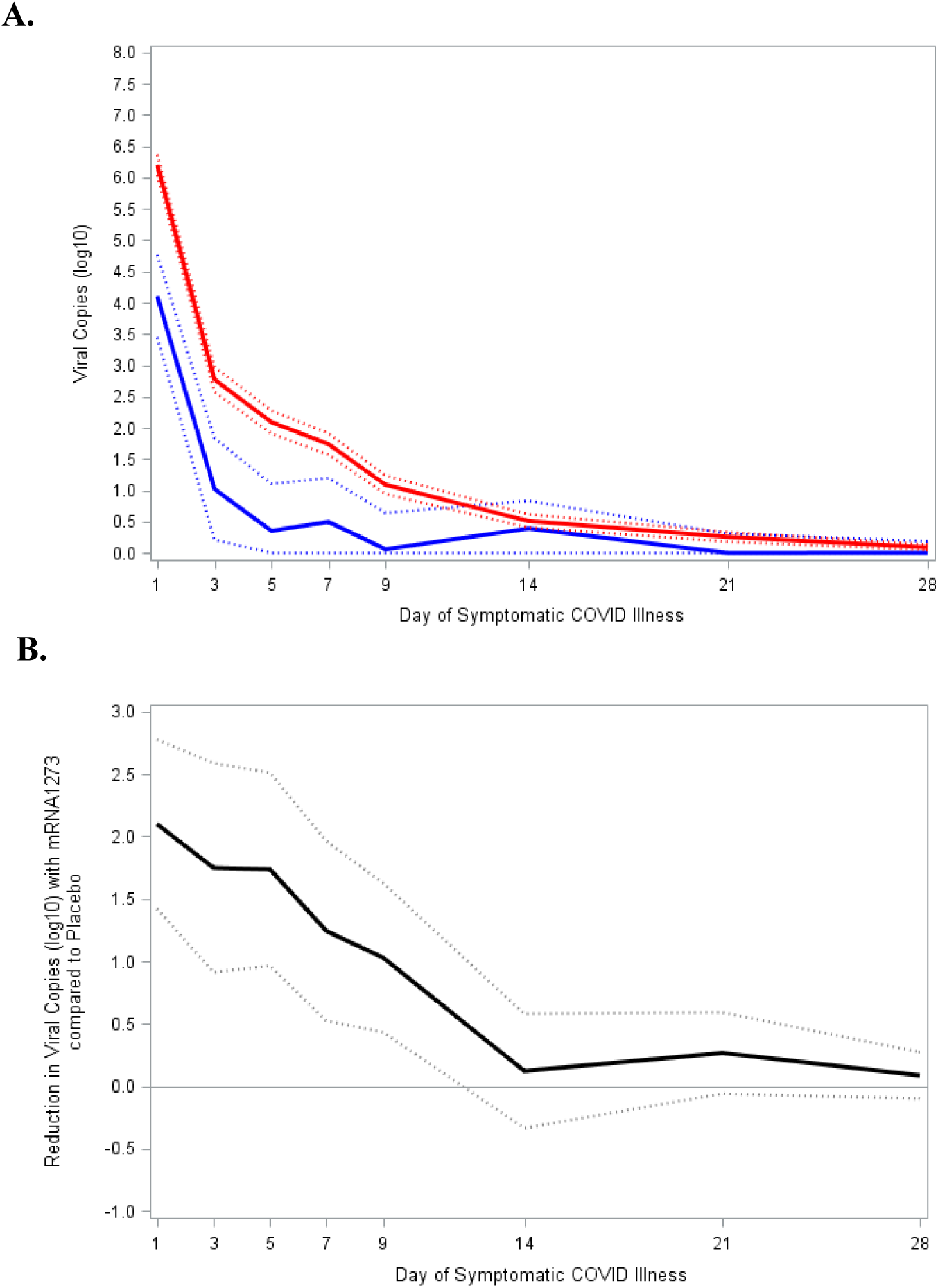
Reduction in SARS-CoV-2 viral load with mRNA-1273 compared with placebo. Viral load was assessed on the basis of SARS-CoV-2 RT-PCR cycle threshold (Ct) values converted to viral copy number as described in the methods. Mixed model repeated measures (MMRM) analysis was performed comparing absolute and change from baseline log10 viral copy between vaccinated and placebo participants based on data from nasopharyngeal swabs at day 1 of illness and saliva samples at days 3, 5, 7, 9, 14, 21, and 28 of illness. Included adjudicated cases in the blinded portion of the study. The mRNA participants (N=36) comprised 29 with first illness and 7 with second illness visits. The placebo participants (N=595) included 527 cases from first illness visits, and 61, 5, and 2 for second third and fourth illness visits respectively. A: Solid lines represent placebo (Red) and mRNA-1273 (Blue), while dotted lines correspondingly denote 95% confidence intervals. B: Difference between the mRNA-1273 and placebo participants in viral copies (log10) black solid line and 95% CI in dotted lines.

### Exploratory Analyses of Burden of Disease (BOD) and Burden of Infection (BOI) in the COVE Trial

Exploratory analyses of BOD and BOI were performed in the COVE trial to assess the effect of vaccination on the severity and symptoms of Covid-19 (Table S5). BOD was assessed in the PP set based on adjudicated cases in participants who had no evidence of SARS-CoV-2 infection by serology and RT-PCR at randomization and had available post-baseline data. The BOD was defined to reflect the severity of symptoms as a set of pre-defined scores (0 [Without Covid-19], 1 [Covid-19 without hospitalization], 2 [Covid-19 with hospitalization, 3 [death]). mRNA-1273 vaccination substantially reduced Covid-19 illness with and without hospitalizations, and Covid-19 deaths. The mean BOD score was higher for the placebo (0.1) than mRNA-1273 (0) group with a resulting VE point estimate for mRNA-1273 on BOD of 93.2% (95% CI 91.0%-94.8%). BOI was evaluated in the PP set based on asymptomatic infections and adjudicated Covid-19 cases in participants who were SARS-CoV-2 infection negative at baseline and had available post-baseline data. The BOI score was similarly defined to that of BOD and additionally included asymptomatic infection (0 [Uninfected], 1/2 [Asymptomatic infection], 1 [Covid-19 without hospitalization], 2 [Covid-19 with hospitalization], 3 [death]). Vaccination reduced asymptomatic infections, as well as Covid-19 with and without hospitalization. The mean BOI score was higher in the placebo (0.06) than mRNA-1273 (0.01) recipients, with a VE of 91.2% (95% CI 89.0%-93.0%). The BOD and BOI scores were similar regardless of age and Covid-19 risk.

### SARS-CoV-2 Variants Detected in the COVE mRNA-1273 Trial

Sequence information was obtained by RT-PCR from nasopharyngeal samples positive for SARS-CoV-2 collected July 2020 through May 2021 from participants with Covid-19 cases in the placebo-controlled portion of the COVE trial which was conducted in the United States.^1^ This included all Covid-19 cases and adjudicated cases. There were a total of 1006 samples available for sequencing (142 for mRNA-1273 and 864 for placebo). Although sequences could not be obtained from all samples, particularly in the mRNA-1273 group as sequencing success diminished with lower copy number, data for the spike gene were generated from 832 different samples (754 placebo and 78 mRNA-1273), corresponding to 791 trial participants (720 from placebo and 71 from mRNA-1273). To assess the relative prevalence of key variant lineages detected in the clinical dataset, Pango lineages^31^ were inferred for each isolate based on amino acid mutations detected in the spike gene (Table S6). Comparison of the prevalence of 589 selected lineages in the clinical study samples with those from a US time-matched subset of the Global Initiative on Sharing All Influenza Data (GISAID) database^30^ revealed that the sequences detected in clinical case samples reflected the circulating strains in the US during the trial, with similar frequencies between the clinical samples and GISAID subset (Figure 3, Tables S7 and S8). In the study dataset, the majority of variants detected were of the B.1/B.1.2 lineage (548 [93%]) and mainly during July 2020 to February 2021. Additionally, variants of the Epsilon (B.1.427/429) lineage (32 [5.4%]), first identified in California, were detected mainly during the months of December 2020 and January 2021, and (2 [1.0%]) Alpha (B1.1.7) variants during March and April (Table S7). Overall, the frequencies of variants corresponding to timeframes of EUA of Covid-19 vaccines and amendment of the COVE study (Dec. 23^rd^, 2020) with transition to the open-label portion (eg January 2021 through May 2021), were also similar to the frequencies in the GISAID database.

**Figure 3.**
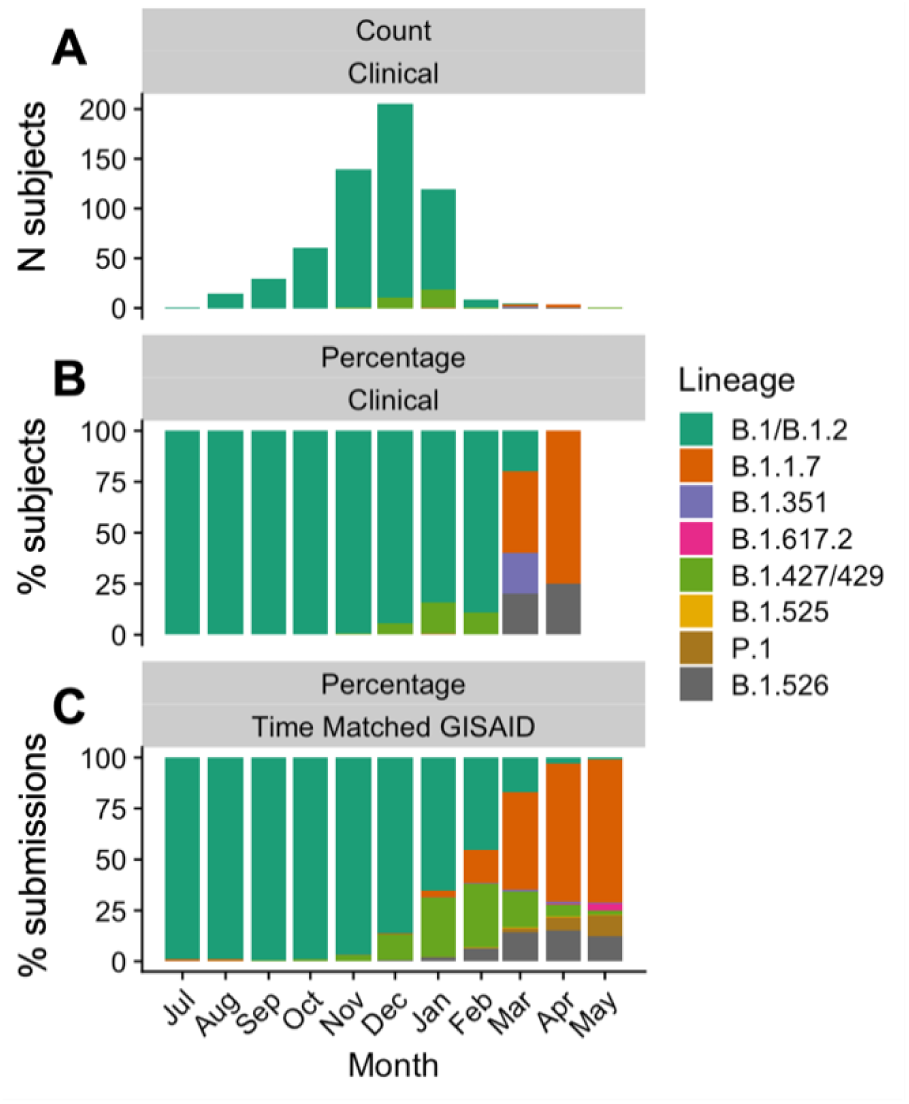
Spike sequence-associated lineages found among COVE trial participants. Summary of selected spike sequence-associated lineages found among COVE trial participants (regardless of symptoms) from July 2020 to May 2021. Y axis: Number of sequences, X axis: Month. A: Number of sequences in the clinical dataset, B: Percent of assigned lineages in the clinical data set, C: Percent of lineages circulating in the US in the same period (time-matched sequence set) obtained from GISAID.^25,30^

### SARS-CoV-2 Variants Among Adjudicated Covid-19 cases in the COVE Trial

Viral variant sequences detected (545 in placebo and 28 in mRNA-1273) among the 825 adjudicated Covid-19 cases (769 in placebo and 56 in mRNA-1273 groups) in the trial starting after randomization in the per-protocol set included 1 (0.1%) of wild-type lineage in the placebo group, with the majority in the B.1.2 (394 [51.2%] and 13 [23.2%]), B.1.243 (23 [3.0%] and 1 [1.8%]) and B.1.596 (13 [1.7%] and 0) lineages in the placebo and mRNA-1273 groups, respectively (Table S9). Overall, 18 (2.3%) of the adjudicated cases starting after randomization were attributed to Epsilon, Gamma and Zeta variants in the placebo group and 3 (5.4%) in the mRNA-1273 group (Tables 1 and S9). Of these, 15 (2.0%) were Epsilon variants first detected in California in the placebo group and 3 (5.4%) in the mRNA-1273 group, including 9 (1.2%) B.1.429 and 6 (0.8%) B.1.427 variants in the placebo and 3 (5.4%) B.1.429 in the mRNA-1273 groups.

**Table 1.**
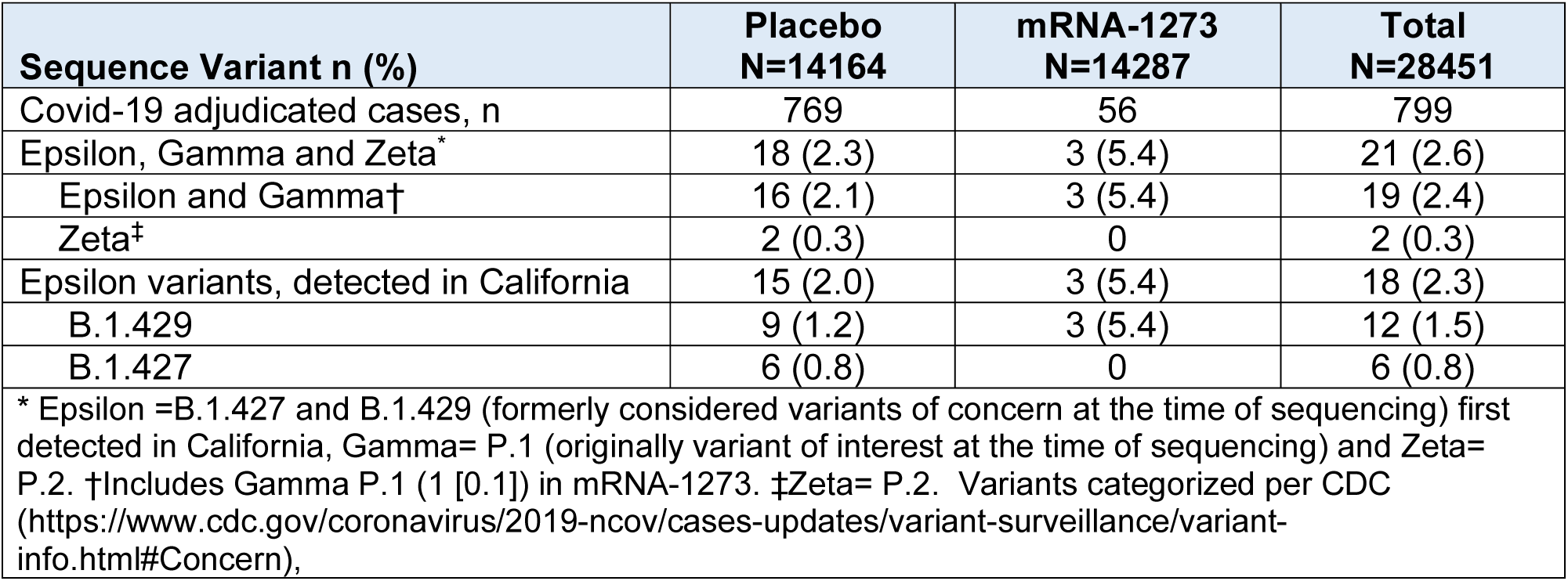
SARS-CoV-2 variants identified in the COVE trial per-protocol set starting after randomization.

Exploratory analyses of blinded phase data (cutoff date March 26, 2021),^2^ were performed to assess the vaccine efficacy (VE) of specific variants in adjudicated Covid-19 cases starting 14 days after the second dose in the PP set. The competing risk method was used, specifically, Fine and Gray’s sub-distribution hazard model, with Covid-19 cases with variants other than the specific variant of interest as competing risk. This exploratory analyses of variant-specific VE was performed for protection against Epsilon variants first detected in California (B.1.427 or B.1.429) as the total number of such cases were >10, and also against the formerly designated VOC and VOI given the interest in these variants.^23,25^ For Epsilon variants first detected in California, the VEs (95% CI) were 81.2% (36.1-94.5%) with 15 cases in the placebo and 3 in the mRNA-1273 groups for combined variants B.1.427 and B.1.429 detected in CA, and 68.9% (−12.8-91.4%; 9 in placebo and 3 in mRNA-1273 groups) for the B.1.429 variant (Table 2). Based on the small number of former VOC (Epsilon, Gamma and Zeta) detected in the placebo and the mRNA-1274 groups (16 and 3 respectively), the VE (95% CI) to prevent Covid-19 of VOC was 82.4% (40.4-94.8%), and the VE was 100.0% (100.0-100.0%) for VOI (2 in placebo and none in the mRNA-1273 groups). Additionally, viral copies in the cases with B.1.427 and B.1.429 variants detected were analyzed using the MMRM, and at least a 10-fold reduction in viral copies (copies/ml log10 [SD]) at the day 1 illness visit for mRNA-1273 (7.4 [0.3]) and placebo (6.0 [0.3]) and shorter duration of detectable viral copies for mRNA-1273 versus placebo was observed (Figure S4), consistent with the viral load reduction of mRNA-1273 seen in overall cases regardless of infecting variant.

**Table 2.** Exploratory analysis of vaccine efficacy against variants in the COVE trial per-protocol set starting 14 days after second dose.

| **Covid-19 Variant Cases in COVE** | **Placebo**<br>**N=14164** | **mRNA-1273**<br>**N=14287** |
| --- | --- | --- |
| Covid-19* primary efficacy endpoint, n (%) | 744 | 55 |
| Vaccine efficacy based on hazard ratio (95% CI)<sup>†</sup> |  | 93.2 (91.0-94.8) |
| Incidence rate per 1,000 person-years (95% CI)<sup>‡§</sup> | 136.6 (127.0-146.8) | 9.6 (7.2-12.5) |
| Covid-19* with Epsilon and Gamma variants, n (%) | 16 (0.1) | 3 (<0.1) |
| Number with competing events, n (%) | 728 (5.1) | 52 (0.4) |
| Vaccine efficacy based on hazard ratio (95% CI)<sup>†</sup> |  | 82.4 (40.4-94.8) |
| Incidence rate per 1,000 person-years (95% CI)<sup>‡§</sup> | 2.9 (1.7-4.8) | 0.52 (0.11-1.5) |
| Covid-19* with Zeta variant, n (%) | 2 (<0.1) | 0 |
| Number with competing events, n (%) | 742 (5.2) | 55 (0.4) |
| Vaccine efficacy based on hazard ratio (95% CI)<sup>†</sup> |  | 100.0 (100.0-100.0) |
| Incidence rate per 1,000 person-years (95% CI)<sup>‡§</sup> | 0.38 (0.04-1.4) | - |
| Covid-19* with Epsilon variants first detected (CA)<sup>¶</sup>, n (%) | 15 (0.1) | 3 (<0.1) |
| Number with competing events, n (%) | 729 (5.1) | 52 (0.4) |
| Vaccine efficacy based on hazard ratio (95% CI)<sup>†</sup> |  | 81.2 (36.1-94.5) |
| Incidence rate per 1,000 person-years (95% CI)<sup>‡§</sup> | 2.8 (1.5-4.5) | 0.52 (0.11-1.5) |
| Covid-19* with Epsilon (B.1.429) variant first detected (CA), n (%) | 9 (0.1) | 3 (<0.1) |
| Number with competing events, n (%) | 735 (5.2) | 52 (0.4) |
| Vaccine efficacy based on hazard ratio (95% CI)<sup>†</sup> |  | 68.9 (-12.8-91.4) |
| Incidence rate per 1,000 person-years (95% CI)<sup>‡§</sup> | 1.7 (0.8-3.2) | 0.52 (0.11-1.6) |
| Variants include Epsilon (B.1.427, B.1.429), Gamma (P.1), and Zeta (P.2). CA=California; CI=confidence interval. *Based on participants with adjudicated assessments starting 14 days after second injection in the per-protocol set, with censoring rules for efficacy analysis. If participant had a positive RT-PCR at the pre-dose 2 visit (day 29) without eligible symptoms with 14 days, or positive (Elecys nucleocapsid [NP]) at scheduled visits prior to becoming a Covid-19 case, the participant was censored at the date of positive RT-PCR or Elecys NP. Covid-19 cases with variant lineages other than the specified variant(s) assessed are considered as competing events. †Vaccine efficacy, defined as 1 - hazard ratio (mRNA-1273 vs. placebo), and 95% CI are estimated using Fine and Gray's sub-distribution hazard model with disease cases as competing events and with the treatment group as a covariate, adjusting for stratification factor. ‡Person-years defined as the total years from randomization date to the earliest of the date of symptomatic SARS-CoV-2 infection, the date of asymptomatic SARS-CoV-2 infection, last date of study participation, or efficacy data cutoff date, whichever was earlier. §Incidence rate is defined as the number of participants with an event divided by the number of participants at risk and adjusted by person-years (total time at risk) in each treatment group. The 95% CI was calculated using the exact method (Poisson distribution) and adjusted by person-years. ¶Includes both Epsilon B.1.427 and B.1.429 variants. Note that variants B.1.427 and B.1.429 were originally categorized as variants of concern (VOC) then reconsidered as variants of interest (VOI), and now as variants being monitored (VBM) by the CDC. Variant Gamma P.1 formerly considered VOC, now VBM. |  |  |

### PCR-Detected Respiratory Pathogens During Symptomatic Study Visits in the COVE Study

The presence of viral and bacterial respiratory pathogens was detected during August 2020 to June 2021 in 954 total nasopharyngeal samples combined from placebo and mRNA-1273 recipients collected at illness visits in parallel with SARS-CoV-2 detection. From the initiation of the study in July 2020 to June 2021, 463 placebo participants had detected respiratory pathogens versus 491 for mRNA-1273 participants. Participant illness visits peaked in December 2020 with 1,002 visits. That month, 461 participants had tested positive for SARS-CoV-2 and/or other respiratory pathogens, with 78% attributed to SARS-CoV-2 and 22% to human rhinoviruses/enteroviruses. Coincident with EUA of Covid-19 vaccines by February 2021, the percentage of SARS-CoV-2 infections (regardless of symptoms) decreased to 39% of all positive results (n=29/74) and by June 2021 to 7% (n=9/126, Figure 4). As the rate of SARS-CoV-2 positivity decreased among participants along with the overall number of respiratory illness visits, the percentage of detection of other respiratory pathogens increased. During August to December 2020, 61% (758) of all positive samples (n=1227) from illness visits were attributed to SARS-CoV-2 infections, and 38% (469) to other respiratory pathogens, mainly rhinovirus/enterovirus infections with 223 in the placebo versus 232 infections in the mRNA-1273 group. From March to June 2021, 85% (365/431) of all positive illness results were attributed to respiratory pathogens other than SARS-COV-2, the diversity of which increased through the spring months. There were 23 cases of co-infection with both SARS-CoV-2 and another respiratory pathogen during October 2020 to June 2021. The majority (19/23) were co-infections with human rhinoviruses/enteroviruses, and there were 3 cases of co-infection with a seasonal Coronavirus strain and 1 with human parainfluenza virus type 3 (HPIV3).

**Figure 4.**
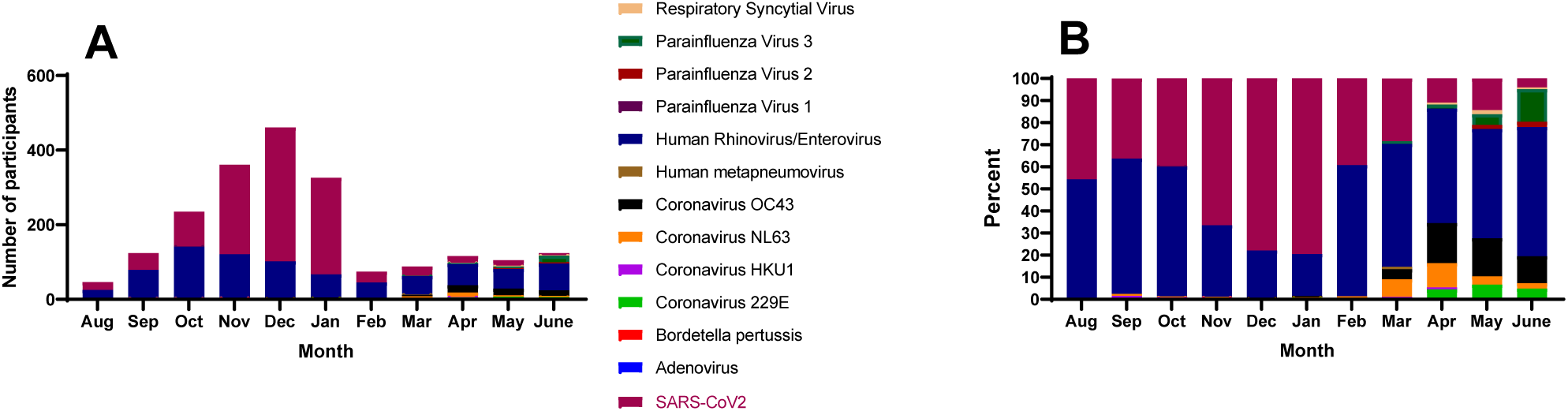
Respiratory pathogens detected in the COVE study per month. Respiratory pathogen sequences detected during August 2020 through June 2021 in samples from all participant illness visits: (A) Number of positive samples and (B) Percent of detected pathogen.

## Discussion

In this initial evaluation of the viral dynamics of SARS-CoV-2 infection in the ongoing COVE mRNA-1273 trial, vaccination with mRNA-1273 significantly reduced the number of SARS-CoV-2 viral copies and duration of shedding in respiratory samples from participants with Covid-19, compared with placebo. Consistent with the effect on viral load, vaccination with mRNA-1273 reduced the severity and symptoms of Covid-19 and SARS-CoV-2 infection in terms of hospitalizations, mortality, and asymptomatic infections based on pre-defined BOD and BOI scores. Sequencing of the SARS-CoV-2 spike gene in clinical samples from the adjudicated cases in both the placebo and mRNA-1273 groups in the trial revealed that variant lineages corresponded to the epidemiological patterns of circulating variants in the US from the start of the study through the blinded and open-label portions of the study. An exploratory analysis of SARS-CoV-2 VOC (Epsilon, Gamma and Zeta) and VOI (Zeta) detected in the placebo and mRNA-1273 groups, resulted in estimated vaccine efficacies (95% CI) of 82.4% (40.4%-94.8%) for VOC, 100% for VOIs and 81.2% (36.1%-94.5%) for Epsilon B.1.427 and B.1.429 variants. Additionally, the majority of symptomatic respiratory cases during the blinded, randomized, portion of the trial, were attributed to SARS-CoV-2 while the percentage of non-SARS-CoV-2 respiratory pathogens increased, coincidently with the availability of Covid-19 vaccines under EUA and cross-over of placebo recipients to mRNA-1273 in the COVE study starting in January 2021.

Studies have shown that higher viral load assessed by Ct values (lower Ct values) and/or converted to copies/ml (as assessed in this study), is related to severe COVID-19 and mortality.^3-10^ Covid-19 vaccination has been shown to attenuate viral load and the duration of viral shedding and illness.^11,12^ In our study, the findings that mRNA-1273 elicited a highly significant reduction in both viral load on the day of Covid-19 illness diagnosis and persistence of viral RNA in saliva samples indicative of viral shedding up to day 9, are relevant as potential surrogates for the transmissibility of the SARS-CoV-2 virus.^4,6,10^ In line with these findings and the efficacy results reported at the end of the blinded part of the COVE trial,^1,2^ mRNA-1273 reduced the symptoms and severity of disease and infection as reflected by the lower BOD and BOI scores seen in vaccine recipients, and is consistent with recent studies showing that vaccination is highly effective in preventing severe Covid-19, asymptomatic infection and Covid-19-related hospitalizations and deaths.^32-35^ Although the transmission dynamics of Covid-19 are still under study, the estimated 10-to 100-fold reductions in viral load, with statistically significant reductions in copies through day 9, may provide a significant reduction in Covid-19 disease spread, even in vaccine breakthrough cases.

The spike gene sequences from the Covid-19 cases in the trial showed that the variants detected were representative of circulating variants in the US during the observation period of the study through June 2021. The high efficacy of the vaccine was originally demonstrated during a period of a relatively lower diversity of circulating strains (July 2020 through December 2020) that corresponded to the bulk of the blinded and placebo-controlled phase of the trial, with data through March 2021, consistent with the high VE for the placebo-controlled part of the trial, and the number of breakthrough cases remained low during the open-label phase. The efficacy data from this exploratory analysis of specific variants of Covid-19 cases, combined with the available respiratory pathogen data through June 2021 suggest that high vaccine efficacy was maintained during the time-frame, which encompassed not only the peak of the US epidemic (January 2021), but also the emergence of Alpha, Beta, Epsilon and Gamma variants. Supported by the sequence data, an exploratory analysis of the VE of mRNA-1273 against circulating variants was performed. Of all VOC, a sufficient number of cases was accrued during the blinded phase to allow for a formal analysis only of the Epsilon variants first detected in California (B.1.427 and B.1.429 variants) with a VE (95% CI) of 81.2% (36.1-94.5) against both California variants B.1.427 and B.1.429. Thus, mRNA-1273 maintained high efficacy against the relatively small number of VOIs and VOCs circulating through April 2021. While mRNA-1273 has remained a highly efficacious vaccine, there is a potential for variants to generate vaccine breakthroughs as more transmissible and divergent variants emerge amidst potentially waning immune responses months after vaccination.^19,23-25,36^

Infection with SARS-CoV-2 causes a broad range of symptoms from asymptomatic or mild symptoms to severe illness including and death; however, co-infections with respiratory pathogens can also contribute to these symptoms, complicating diagnosis and patient management.^27-29^ The finding that the majority of symptomatic respiratory cases were caused by SARS-CoV-2 infection and mostly among placebo recipients during the blinded portion of the study further confirms the high efficacy of mRNA-1273 vaccine in the prevention of symptomatic, molecularly confirmed, Covid-19 disease.^1,2^ The percentage drop in SARS-CoV-2 infection and increase in the percentage of non-SARS-CoV-2 respiratory pathogens among trial participants during February 2021 and through June 2021 when participants in the placebo group began crossing over to receive mRNA-1273 is consistent with the high efficacy of the vaccine against SARS-CoV-2 at a time when relaxing infection prevention measures resulted in the emergence of other respiratory viruses. The presence of other respiratory viruses was similar between the mRNA-1273 and placebo groups, indicating that the case ascertainment between the 2 groups was unbiased, and also not impacted by mRNA-1273 vaccination. The observations of high efficacy combined with a reduced viral load in breakthrough cases and reduced burden of disease and infections demonstrate that immunization with mRNA-1273 does not lead to vaccine enhanced respiratory disease, a theoretical concern at the onset of the Covid vaccine development.^1,2^

There are several limitations to consider. First, it should be noted that the analyses presented here are based upon an initial evaluation of data that will continue to accrue over time and will be subject to future updates. Although the BOD and BOI results were statistically significant over the entire PP set, the results were driven by the overall high vaccine efficacy and a numerically dominant prevalence of mild-to-moderate disease, including asymptomatic infections for BOI. With increased follow-up times and inevitably acquired disease in participants, the proportions of BOD and BOI may change. A prespecified, proportional means model was used for the analysis, to allow direct comparison across different periods of follow-up, minimizing these effects. The sequence data also come with known caveats, some demonstrated through this work. For example, the fact that the vaccine has a marked effect on lowering SARS-CoV-2 viral loads hampered our efforts to generate an unbiased sequence data set. Even though sequencing efforts were performed in a blinded manner and the team was unaware of participant treatment assignments, the success rates in obtaining good quality sequences from samples of vaccine breakthrough cases was 50% or less, while it was over 80% for placebo-originated samples. Although this is a clear, unavoidable bias in the available sequence data, it does not affect the overall assessment of the biological effect of vaccination on viral load. While a highly-aggressive variant capable of causing both a higher number of cases and viral loads among vaccine recipients was not previously seen, the emergence of the delta variant may yield samples that can be more readily sequenced and hence, a resulting detection bias in its favor.^37,38^ Finally, we must also recognize that the small sample size of variants detected limits the assessment and interpretation of VE against emerging variants. A further limitation is that the treatment groups in the original 1:1 randomized blinded and placebo-controlled trial had evolved, as an increasing number of placebo recipients either crossed over to be immunized or left the study to seek vaccination outside of the study.

In summary, this analysis of the viral load and the circulating viral variants in the mRNA-1273 phase 3 COVE trial during the placebo-controlled phase suggests that vaccination with mRNA-1273 vaccine leads to a significant reduction of the SARS-CoV-2 viral load and shedding period, and associated BOD and BOI. mRNA-1273 vaccine showed high efficacy and continues to have a marked effect in reducing symptomatic Covid-19 cases among vaccine recipients. The shifting landscape in terms of new emerging variants and the potential waning of the immune response suggest that a continuation of these studies is warranted.

## Supporting information

Consort checklist

Protocol

## Data Availability

As the trial is ongoing, access to patient-level data and supporting clinical documents with qualified external researchers may be available upon request and subject to review once the trial is complete.

https://www.ncbi.nlm.nih.gov/pubmed/33378609

https://www.ncbi.nlm.nih.gov/pubmed/34551225

## Author Contributions

Drs. Pajon, Zhou, Leav, Kacena, Schodel, Baden, El Sahly, Essink and Tomassini contributed to the design of the analysis, and Drs. Kacena, Pajon, Zhou, Deng, Girard, Paila, Dixon, Leav, Schodel and Tomassini, and Mr. Zhao and Ms Ding contributed to the analysis and/or interpretation of the data. Drs. Baden, El Sahly, Essink, Mullane, Frank, Denhan, and Kerwin participated in the COVE study investigation, clinical trial data acquisition, and data analysis/interpretation in relation to the manuscript. Drs. Pajon, Kacena, Girard, Paila and Tomassini developed the initial draft. All authors contributed to the review and editing of the manuscript and approved the final version for submission.

