## Supplement for "Initial Analysis of Viral Dynamics and Circulating Viral Variants During the mRNA-1273 Phase 3 COVE Trial"

**Pajon, R. et al.**

### Supplement

#### Table S1. Baseline and demographics characteristics viral copy analysis population

| Characteristics n (%) | Placebo N=653 | mRNA-1273 N=48 |
|---|---|---|
| Age (y) | | |
| Mean (SD) | 48.0 (14.4) | 49.5 (14.6) |
| Median (Min, Max} | 48 (18-87) | 49 (24-74) |
| Sex | | |
| Female | 324 (49.7) | 23 (47.9) |
| Male | 329 (50.3) | 25 (52.1) |
| Race | | |
| White | 560 (85.6) | 43 (89.6) |
| Black or African American | 30 (4.6) | 2 (4.2) |
| Asian | 26 (4.0) | 1 (2.1) |
| American Indian or Alaska Native | 4 (0.6) | 0 |
| Multiple | 8 (1.2) | 1 (2.1) |
| Other | 15 (2.3) | 1 |
| Not reported or unknown | 10 (1.5) | 0 |
| Ethnicity | | |
| Hispanic or Latino | 150 (22.8) | 9 (18.8) |
| Not Hispanic or Latino | 500 (76.8) | 39 (81.3) |
| Not reported or unknown | 3 (0.5) | 0 |
| Risk Factor for Severe Covid-19 at Screening* | | |
| Chronic lung disease | 24 (3.7) | 4 (8.3) |
| Significant cardiac disease | 29 (4.5) | 3 (6.3) |
| Severe obesity | 65 (9.9) | 7 (14.6) |
| Diabetes | 64 (9.8) | 3 (6.3) |
| Liver disease | 5 (0.8) | 1 (2.1) |
| HIV | 2 (0.3) | 0 |
| Body Mass Index, (kg/m$^2$) | | |
| n | 650 | 48 |
| Mean (SD) | 30.4 (7.0) | 32.3 (7.1) |

*Participants could be under one or more categories and were counted once at each category

**Table S2. Viral load (log10) median copies**

| | Viral Load Median (Log10 viral copies/ml) | | |
|---|---|---|---|
| **Illness day** | **Placebo** | **mRNA-1273** | **Difference** |
| 1 | 6.7 | 3.4 | 3.4 |
| 3 | 3.0 | 0 | 3.0 |
| 5 | 2.3 | 0 | 2.3 |
| 7 | 0 | 0 | 0 |
| 9 | 0 | 0 | 0 |
| 14 | 0 | 0 | 0 |
| 21 | 0 | 0 | 0 |
| 28 | 0 | 0 | 0 |

**Table S3. Mixed model repeated measures estimates displayed in Figure 2A**

| | mRNA-1273 (n=36) | | | Placebo (n=595) | | |
|---|---|---|---|---|---|---|
| **Illness Day** | **Estimate of Viral Copies (log10)** | **Lower Bound 95% CI** | **Upper Bound 95% CI** | **Estimate of Viral Copies (log10)** | **Lower Bound 95% CI** | **Upper Bound 95% CI** |
| 1 | 4.10 | 3.44 | 4.76 | 6.20 | 6.04 | 6.37 |
| 3 | 1.02 | 0.21 | 1.84 | 2.77 | 2.58 | 2.97 |
| 5 | 0.35 | 0 | 1.10 | 2.09 | 1.91 | 2.27 |
| 7 | 0.50 | 0 | 1.20 | 1.74 | 1.57 | 1.91 |
| 9 | 0.06 | 0 | 0.64 | 1.09 | 0.94 | 1.24 |
| 14 | 0.39 | 0 | 0.83 | 0.51 | 0.40 | 0.62 |
| 21 | 0.00 | 0 | 0.31 | 0.25 | 0.18 | 0.33 |
| 28 | 0.00 | 0 | 0.18 | 0.09 | 0.05 | 0.13 |

Note: Of the 48 mRNA-1273 viral copy participants, 9 did not have the day 1 illness nasopharyngeal swab available, and another 3 participants had a nasopharyngeal day 1 swab that was negative, leaving 36 for the MMRM modeling. Of the 653 mRNA-1273 viral copy participants, 43 did not have the day 1 illness nasopharyngeal swab available, and another 15 participants had a nasopharyngeal day 1 swab that was negative, leaving 595 for the MMRM modeling.

**Table S4. Viral copy reduction (log10) in mRNA-1273 compared to placebo based on the mixed model repeated measures model Figure 2B**

| Illness Day | Viral copy reduction (Log10) in mRNA-1273 compared with placebo | Lower Bound 95% CI | Upper Bound 95% CI | p-value |
|---|---|---|---|---|
| 1 | 2.10 | 1.42 | 2.78 | <.0001 |
| 3 | 1.75 | 0.91 | 2.59 | <.0001 |
| 5 | 1.74 | 0.96 | 2.51 | <.0001 |
| 7 | 1.24 | 0.52 | 1.96 | 0.0007 |
| 9 | 1.03 | 0.43 | 1.63 | 0.0008 |
| 14 | 0.12 | -0.34 | 0.58 | 0.0616 |
| 21 | 0.27 | -0.06 | 0.59 | 0.1106 |
| 28 | 0.09 | -0.10 | 0.27 | 0.3643 |

### Table S5. Vaccine effect on BOD and BOI

| | Placebo | mRNA-1273 |
|---|---|---|
| | N=14164 | N=14287 |
| n (%) | | |
| Number of participants with each level of BOD score* | | |
| 0 Without Covid-19 | 13394 (94.6) | 14230 (99.6) |
| 1 Covid-19 without hospitalization | 741 (5.2) | 55 (0.4) |
| 2 Covid-19 with hospitalization | 28 (0.2) | 2 (<0.1) |
| 3 Deaths | 1 (<0.1) | 0 |
| Mean of BOD Score (SD)† | 0.1 (0.24) | 0 (0.07) |
| Treatment comparison (mRNA-1273 vs Placebo)‡ | | |
| Ratio of the mean BOD score (95% CI) | | 0.07 (0.05-0.09) |
| 1 - ratio of the mean BOD score (95% CI) | | 93.2 (91.0-94.8) |
| Number of participants with each level of BOI score n (%)§ | | |
| 0 No infection | 13295 (93.9) | 14187 (99.3) |
| 1/2 Asymptomatic infection | 99 (0.7) | 43 (0.3) |
| 1 Covid-19 without hospitalization | 741 (5.2) | 55 (0.4) |
| 2 Covid-19 with hospitalization | 28 (0.2) | 2 (<0.1) |
| 3 Deaths | 1 (<0.1) | 0 |
| Mean of BOI Score (SD) † | 0.06 | 0.01 |
| Treatment comparison (mRNA-1273 vs Placebo)‡ | | |
| Ratio of the mean BOI score (95% CI) | | 0.088 (0.07-0.110) |
| 1 - ratio of the mean BOI score (95% CI) | | 91.2 (89-93) |

BOD = Burden of Disease. BOI = Burden of Infection. BOD was defined based on adjudicated Covid-19 cases. *Number of participants who are without Covid-19 at randomization and have available post-baseline data. Percentages are based on number of participants with BOD score. †The maximum BOD/BOI score from randomization through study completion. ‡Vaccine efficacy is estimated using proportional means model including vaccination group as fixed effect and stratified with stratification factor at randomization. BOI was defined based on asymptomatic infections and adjudicated Covid-19 cases. BOI score of 0=No infection, 1/2=Asymptomatic infection, 1=Covid-19 without hospitalization, 2=COVID-19 with hospitalization, and 3=Death. §Number of participants who are without SARS-CoV-2 infection at baseline and have available post-baseline data. Percentages are based on number of participants with BOI score.

### Table S6. Variants classification (as of September 30, 2021)[1]

| Variant (PANGO)† | Spike protein substitutions | Name (Nextstrain)‡ | First Detected | WHO Label | Original designations |
|---|---|---|---|---|---|
| **VBM** | | | | | |
| B.1.1.7 | Δ69/70, Δ144, (E484K*) (S494P*), N501Y, A570D, D614G, P681H, T716I, S982A, D1118H (K1191N*) | 20I/501Y.V1 | United Kingdom | Alpha | VOC |
| B.1.351, B.1.351.2, B.1.351.3 | D80A, D215G, Δ241/242/243, K417N, E484K, N501Y, D614G, A701V | 20H/501.V2 | South Africa | Beta | VOC |
| P.1, P.1.1, P.1.2 | L18F, T20N, P26S, D138Y, R190S, K417T, E484K, N501Y, D614G, H655Y,T1027I | 20J/501Y.V3 | Japan/Brazil | Gamma | VOC |
| B.1.427§ | L452R, D614G | 20C/S:452R | United States-(California) | Epsilon | VOC, VOI |
| B.1.429§ | S13I, W152C, L452R, D614G | 20C/S:452R | United States-(California) | Epsilon | VOC, VOI |
| B.1.525 | A67V, 69del, 70del, 144del, E484K,D614G, Q677H, F888L | 20A/S:484K | United Kingdom/Nigeria December 2020 | Eta | VOI |
| B.1.526 | L5F, (D80G*), T95I, (Y144*), (F157S*), D253G, (L452R*), (S477N*), E484K, D614G, A701V, (T859N*), (D950H*), (Q957R*) | 20C/S:484K | United States (New York) – November 2020 | Iota | VOI |
| B.1.617.1 | (T95I), G142D, E154K, L452R, E484Q, D614G, P681R, Q1071H | 20A/S:154K | India – December 2020 | Kappa | VOI |
| B.1.617.3 | T19R, G142D, L452R, E484Q, D614G, P681R, D950N | 20A | India – October 2020 | None | VOI |
| B.1.621 | T95I, Y144S, Y145N, R346K, E484K or the escape mutation, N501Y, D614G, P681H, and D950N. | | India | Mu | VBM |
| P.2 | E484K, D614G, and V1176F, F565L* | | India | Zeta | VOI |
| **VOC** | | | | | |
| B.1.617.2, AY.1, AY.2, AY.3, AY.4, AY.5, AY.6, AY.7, AY.8, Ay.9, AY.10, AY.11, AY.12 | T19R, (V70F*), T95I, G142D, E156-, F157-, R158G, (A222V*), (W258L*), (K417N*), L452R, T478K, D614G, P681R, D950N | 21A/S:478K | India | Delta | VOC |

VBM=Variant being monitored. VOC=Variant of concern. VOI=Variant of interest. *detected in some sequences but not all. †PANGO (Phylogenetic Assignment of Named Global Outbreak) nomenclature of lineages. PANGO is a software tool developed by members of the Rambaut Lab, and associated web application was developed by the Centre for Genomic Pathogen Surveillance in South Cambridgeshire, intended to implement the dynamic nomenclature of SARS-CoV-2 lineages. Next strain nomenclature. ‡Nextstrain collaboration between researchers in Seattle, USA and Basel, Switzerland, provides open-source tools for visualizing the genetics of outbreaks with a goal to support public health surveillance by facilitating understanding of the spread and evolution of pathogens. §Initially categorized as VOC, then a VOI and currently no longer considered a VOI. Source: CDC https://www.cdc.gov/coronavirus/2019-ncov/cases-updates/variant-surveillance/variant-info.html#Concern

**Variant Being Monitored (VBM):** Variants being monitored for which there are data indicating a potential or clear impact on approved or authorized medical countermeasures or that has been associated with more severe disease or increased transmission but are no longer detected or are circulating at very low levels in the United States, and as such, do not pose a significant and imminent risk to public health in the United States.
**Variants of Interest (VOI):** A variant with specific genetic markers that have been associated with changes to receptor binding, reduced neutralization by antibodies generated against previous infection or vaccination, reduced efficacy of treatments, potential diagnostic impact, or predicted increase in transmissibility or disease severity.
**Variants of Concern (VOC):** A variant for which there is evidence of an increase in transmissibility, more severe disease (increased hospitalizations or deaths), significant reduction in neutralization by antibodies generated during previous infection or vaccination, reduced effectiveness of treatments or vaccines, or diagnostic detection failures.
**Variants of High Consequence:** A variant of high consequence has clear evidence that prevention measures or medical countermeasures (MCMs) have significantly reduced effectiveness relative to previously circulating variants.

### Table S7. Counts for select lineages

| Revised lineage | 2020 | | | | | | 2021 | | | | |
|---|---|---|---|---|---|---|---|---|---|---|---|
| | Jul | Aug | Sept | Oct | Nov | Dec | Jan | Feb | March | April | May |
| **Time-matched GISAID** | | | | | | | | | | | |
| B.1/B.1.2 | 3290 | 3336 | 2742 | 4468 | 11986 | 15442 | 27287 | 20756 | 15842 | 3984 | 580 |
| B.1.1.7 | 29 | 37 | 0 | 1 | 19 | 136 | 1472 | 7386 | 44563 | 92057 | 49674 |
| B.1.351 | 0 | 0 | 0 | 0 | 0 | 0 | 36 | 157 | 626 | 989 | 484 |
| B.1.617.2 | 2 | 0 | 0 | 0 | 0 | 1 | 5 | 8 | 20 | 765 | 2459 |
| B.1.427/429 | 4 | 4 | 18 | 36 | 375 | 2263 | 12253 | 14245 | 16612 | 8137 | 1523 |
| B.1.525 | 0 | 0 | 0 | 0 | 0 | 1 | 32 | 86 | 396 | 498 | 172 |
| P.1 | 0 | 0 | 0 | 0 | 0 | 0 | 10 | 49 | 1674 | 8586 | 7307 |
| B.1.526 | 1 | 0 | 1 | 3 | 4 | 84 | 779 | 2920 | 13258 | 20514 | 8564 |
| **Clinical study samples** | | | | | | | | | | | |
| B.1/B.1.2 | 1 | 15 | 30 | 61 | 138 | 194 | 100 | 8 | 1 | 0 | 0 |
| B.1.1.7 | 0 | 0 | 0 | 0 | 0 | 0 | 0 | 0 | 2 | 3 | 0 |
| B.1.351 | 0 | 0 | 0 | 0 | 0 | 0 | 0 | 0 | 1 | 0 | 0 |
| B.1.617.2 | 0 | 0 | 0 | 0 | 0 | 0 | 0 | 0 | 0 | 0 | 0 |
| B.1.427/429 | 0 | 0 | 0 | 0 | 1 | 11 | 18 | 1 | 0 | 0 | 1 |
| B.1.525 | 0 | 0 | 0 | 0 | 0 | 0 | 0 | 0 | 0 | 0 | 0 |
| P.1 | 0 | 0 | 0 | 0 | 0 | 0 | 1 | 0 | 0 | 0 | 0 |
| B.1.526 | 0 | 0 | 0 | 0 | 0 | 0 | 0 | 0 | 1 | 1 | 0 |

### Table S8. Percentage for select lineages time-matched GISAID and clinical datasets

| Revised lineage | 2020 | | | | | | 2021 | | | | |
|---|---|---|---|---|---|---|---|---|---|---|---|
| | Jul | Aug | Sept | Oct | Nov | Dec | Jan | Feb | March | April | May |
| **Time-matched GISAID** | | | | | | | | | | | |
| B.1/B.1.2 | 98.9 | 98.8 | 99.3 | 99.1 | 96.8 | 86.1 | 65.2 | 45.5 | 17.0 | 2.9 | 0.8 |
| B.1.1.7 | 0.9 | 1.1 | 0.0 | 0.0 | 0.2 | 0.8 | 3.5 | 16.2 | 47.9 | 67.9 | 70.2 |
| B.1.351 | 0.0 | 0.0 | 0.0 | 0.0 | 0.0 | 0.0 | 0.1 | 0.3 | 0.7 | 0.7 | 0.7 |
| B.1.617.2 | 0.1 | 0.0 | 0.0 | 0.0 | 0.0 | 0.0 | 0.0 | 0.0 | 0.0 | 0.6 | 3.5 |
| B.1.427/429 | 0.1 | 0.1 | 0.7 | 0.8 | 3.0 | 12.6 | 29.3 | 31.2 | 17.9 | 6.0 | 2.2 |
| B.1.525 | 0.0 | 0.0 | 0.0 | 0.0 | 0.0 | 0.0 | 0.1 | 0.2 | 0.4 | 0.4 | 0.2 |
| P.1 | 0.0 | 0.0 | 0.0 | 0.0 | 0.0 | 0.0 | 0.0 | 0.1 | 1.8 | 6.3 | 10.3 |
| B.1.526 | 0.0 | 0.0 | 0.0 | 0.1 | 0.0 | 0.5 | 1.9 | 6.4 | 14.3 | 15.1 | 12.1 |
| **Clinical study samples** | | | | | | | | | | | |
| B.1/B.1.2 | 100.0 | 100.0 | 100.0 | 100.0 | 99.3 | 94.6 | 84.0 | 88.9 | 20.0 | 0.0 | 0.0 |
| B.1.1.7 | 0.0 | 0.0 | 0.0 | 0.0 | 0.0 | 0.0 | 0.0 | 0.0 | 40.0 | 75.0 | 0.0 |
| B.1.351 | 0.0 | 0.0 | 0.0 | 0.0 | 0.0 | 0.0 | 0.0 | 0.0 | 20.0 | 0.0 | 0.0 |
| B.1.617.2 | 0.0 | 0.0 | 0.0 | 0.0 | 0.0 | 0.0 | 0.0 | 0.0 | 0.0 | 0.0 | 0.0 |
| B.1.427/429 | 0.0 | 0.0 | 0.0 | 0.0 | 0.7 | 5.4 | 15.1 | 11.1 | 0.0 | 0.0 | 100.0 |
| B.1.525 | 0.0 | 0.0 | 0.0 | 0.0 | 0.0 | 0.0 | 0.0 | 0.0 | 0.0 | 0.0 | 0.0 |
| P.1 | 0.0 | 0.0 | 0.0 | 0.0 | 0.0 | 0.0 | 0.8 | 0.0 | 0.0 | 0.0 | 0.0 |
| B.1.526 | 0.0 | 0.0 | 0.0 | 0.0 | 0.0 | 0.0 | 0.0 | 0.0 | 20.0 | 25.0 | 0.0 |

### Table S9. Summary of variants in adjudicated Covid-19 cases starting after randomization in the per-protocol set

| Covid-19 n (%) | Placebo<br>N=14164 | mRNA-1273<br>N=14287 |
|---|---|---|
| COVID-19* cases, n (%) | 769 (5.4) | 56 (0.4) |
| Number of events by lineage, n (%) | | |
| B.1 | 5 (0) | - |
| B.1.1 | 1 (0) | - |
| B.1.1.128 | 1 (0) | - |
| B.1.1.186 | 2 (0) | - |
| B.1.1.207 | 1 (0) | - |
| B.1.1.222 | 8 (0.1) | - |
| B.1.1.316 | 1 (0 | - |
| B.1.1.337 | 1 (0) | - |
| B.1.1.432 | 1 (0) | - |
| B.1.1.434 | 1 (0) | - |
| B.1.1.519 | 2 (0) | - |
| B.1.2 | 394 (2.8) | 13 (0.1) |
| B.1.232 | 1 (0) | - |
| B.1.234 | 6 (0) | - |
| B.1.240 | 1 (0) | - |
| B.1.243 | 23 (0.2) | 1 (0) |
| B.1.311 | 6 (0) | - |
| B.1.349 | 1 (<0.0) | - |
| B.1.369 | 2 (<0.0) | - |
| B.1.375 | 1 (<0.0) | - |
| B.1.382 | 1 (<0.0) | - |
| B.1.396 | 1 (<0.0) | - |
| B.1.404 | 2 (<0.0) | - |
| B.1.427 | 6 (<0.0) | - |
| B.1.429 | 9 (0.1) | 3 (<0.0) |
| B.1.517 | 2 (<0.0) | - |
| B.1.526.3 | 1 (<0.0) | - |
| B.1.544 | 2 (<0.0) | - |
| B.1.551 | 1 (<0.0) | - |
| B.1.561 | 5 (<0.0) | - |
| B.1.564 | 3 (<0.0) | - |
| B.1.587 | 8 (0.1) | - |
| B.1.595 | 2 (<0.0) | - |
| B.1.596 | 13 (0.1) | - |
| B.1.599 | 1 (<0.0) | - |
| B.1.605 | 1 (<0.0) | - |
| B.1.609 | 1 (0) | - |
| NONE | 20 (0.1) | - |
| P.1 | 1 (0) | - |
| P.2 | 2 (0) | - |
| R.1 | 3 (0) | - |
| WILD TYPE | 1 (0) | - |
| By First Detected, n (%) | | |
| Brazil | 1 (0) | - |
| P.1 | 1 (0) | - |
| California | 15 (0.1) | 3 (0) |
| B.1.427 | 6 (0) | |
| B.1.429 | 9 (0.1) | 3 (0) |
| Variant of Concern | | |
| B.1.427 | 6 (0) | |
| B.1.429 | 9 (0.1) | 3 (0) |
| P.1 | 1 (0) | |
| Variant of Interest | | |
| P.2 | 2 (0) | |

NONE=unable to assign to a lineage. *With the censoring rules for efficacy analyses. COVID-19 case is based on eligible symptoms and positive RT-PCR within 14 days. Percentages based on adjudicated cases starting 14 days post-randomization in the per-protocol set. If a participant had a positive RT-PCR test at pre-dose 2 visit (day 29) without eligible symptoms with 14 days, or positive Elecsys at scheduled visits prior to becoming a COVID-19 case, the participant was censored at the date with positive RT-PCR or Elecsys. Table 14.2.1.1.2.1.4.1

### Figure S1: SARS-CoV-2 circulating in the US Jan-Mar 2021

#### A: Jan-Mar 2021

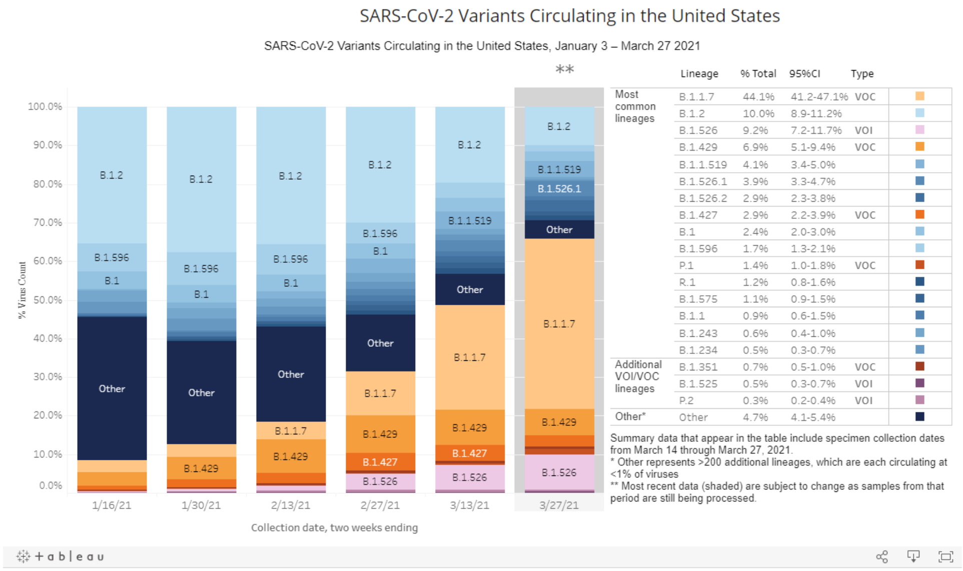

| | Lineage | % Total | 95%CI | Type |
|---|---|---|---|---|
| Most common lineages | B.1.1.7 | 44.1% | 41.2-47.1% | VOC |
| | B.1.2 | 10.0% | 8.9-11.2% | |
| | B.1.526 | 9.2% | 7.2-11.7% | VOI |
| | B.1.429 | 6.9% | 5.1-9.4% | VOC |
| | B.1.1.519 | 4.1% | 3.4-5.0% | |
| | B.1.526.1 | 3.9% | 3.3-4.7% | |
| | B.1.526.2 | 2.9% | 2.3-3.8% | |
| | B.1.427 | 2.9% | 2.2-3.9% | VOC |
| | B.1 | 2.4% | 2.0-3.0% | |
| | B.1.596 | 1.7% | 1.3-2.1% | |
| | P.1 | 1.4% | 1.0-1.8% | VOC |
| | R.1 | 1.2% | 0.8-1.6% | |
| | B.1.575 | 1.1% | 0.9-1.5% | |
| | B.1.1 | 0.9% | 0.6-1.5% | |
| | B.1.243 | 0.6% | 0.4-1.0% | |
| | B.1.234 | 0.5% | 0.3-0.7% | |
| Additional VOI/VOC lineages | B.1.351 | 0.7% | 0.5-1.0% | VOC |
| | B.1.525 | 0.5% | 0.3-0.7% | VOI |
| | P.2 | 0.3% | 0.2-0.4% | VOI |
| Other* | Other | 4.7% | 4.1-5.4% | |

† Estimated weights come from laboratory data providing the number of reverse transcription polymerase chain reaction (RT-PCR) tests and number of positive RT-PCR test results stratified by state, specimen collection date, and by genomic surveillance data source, using a survey-design-based approach. COVID-19 laboratory data sources include commercial and reference laboratories, public health laboratories, hospital laboratories, and other testing locations

#### B: Mar-Jun 2021

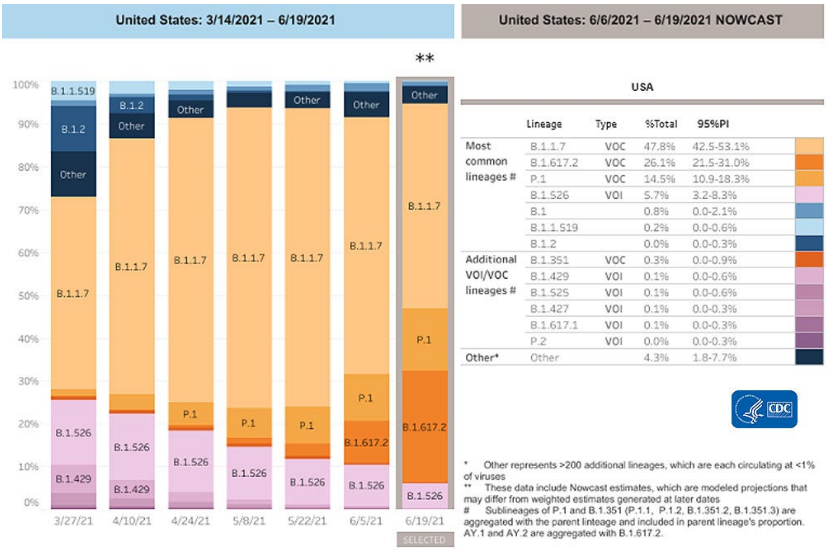

| | Lineage | Type | %Total | 95%PI |
|---|---|---|---|---|
| Most common lineages # | B.1.1.7 | VOC | 47.8% | 42.5-53.1% |
| | B.1.617.2 | VOC | 26.1% | 21.5-31.0% |
| | P.1 | VOC | 14.5% | 10.9-18.3% |
| | B.1.526 | VOI | 5.7% | 3.2-8.3% |
| | B.1 | | 0.8% | 0.0-2.1% |
| | B.1.1.519 | | 0.2% | 0.0-0.6% |
| | B.1.2 | | 0.0% | 0.0-0.3% |
| Additional VOI/VOC lineages # | B.1.351 | VOC | 0.3% | 0.0-0.9% |
| | B.1.429 | VOI | 0.1% | 0.0-0.6% |
| | B.1.525 | VOI | 0.1% | 0.0-0.6% |
| | B.1.427 | VOI | 0.1% | 0.0-0.3% |
| | B.1.617.1 | VOI | 0.1% | 0.0-0.3% |
| | P.2 | VOI | 0.0% | 0.0-0.3% |
| Other* | Other | | 4.3% | 1.8-7.7% |

**June 2021 estimates of SARS-CoV-2 variants circulating in the US Prevalence**
The data below shows the estimated biweekly prevalence of the most common SARS-CoV-2 lineages circulating in the United States, based on >40,000 sequences collected through CDC's national genomic surveillance since Dec 20, 2020 and grouped in 2-week intervals. Variant proportions in Figure 1 are adjusted using statistical weighting† to correct for the non-random sampling of sequencing data over time and across states and to provide more representative national estimates

(source: https://covid.cdc.gov/covid-data-tracker/?CDC_AA_refVal=https%3A%2F%2Fwww.cdc.gov%2Fcoronavirus%2F2019-ncov%2Fcases-updates%2Fvariant-proportions.html#variant-proportions)

**Figure S2. Viral copy number by age groups ≥18-<65 and ≥65 years**

**A. ≥18-<65**

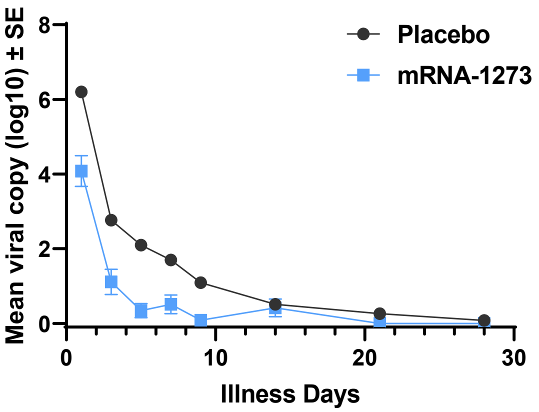

**B. ≥65**

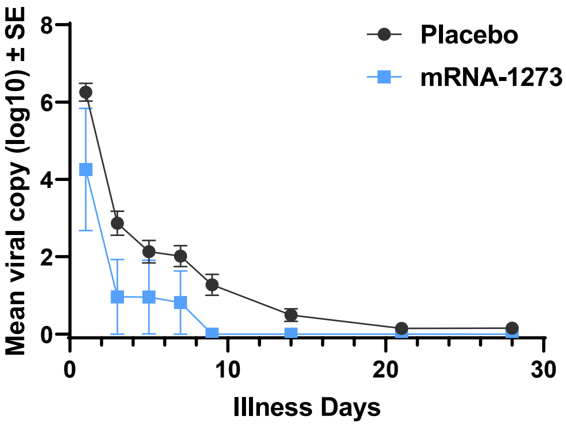

Data underlying graphs above.

| Illness Day | ≥18-<65 years | | | ≥65 years | | |
|---|---|---|---|---|---|---|
| | n | Log10 viral copies | | n | Log10 viral copies | |
| | | Mean | Std Error | | Mean | Std Error |
| Placebo | | | | | | |
| 1 | 514 | 6.20 | 0.09 | 81 | 6.25 | 0.23 |
| 3 | 442 | 2.76 | 0.11 | 63 | 2.87 | 0.31 |
| 5 | 440 | 2.10 | 0.10 | 61 | 2.13 | 0.30 |
| 7 | 450 | 1.70 | 0.10 | 64 | 2.02 | 0.27 |
| 9 | 411 | 1.09 | 0.08 | 55 | 1.27 | 0.27 |
| 14 | 407 | 0.51 | 0.06 | 62 | 0.49 | 0.16 |
| 21 | 398 | 0.26 | 0.04 | 55 | 0.15 | 0.10 |
| 28 | 408 | 0.08 | 0.02 | 66 | 0.15 | 0.09 |
| mRNA-1273 | | | | | | |
| 1 | 32 | 4.08 | 0.41 | 4 | 4.26 | 1.58 |
| 3 | 27 | 1.11 | 0.34 | 3 | 0.96 | 0.96 |
| 5 | 26 | 0.34 | 0.19 | 4 | 0.95 | 0.95 |
| 7 | 26 | 0.51 | 0.25 | 4 | 0.82 | 0.82 |
| 9 | 28 | 0.08 | 0.08 | 4 | 0 | 0 |
| 14 | 25 | 0.42 | 0.24 | 1 | 0 | - |
| 21 | 23 | 0.00 | 0.00 | 3 | 0 | 0 |
| 28 | 24 | 0.00 | 0.00 | 3 | 0 | 0 |

**Figure S3. Time to undetectable SARS-CoV-2 viral copies**

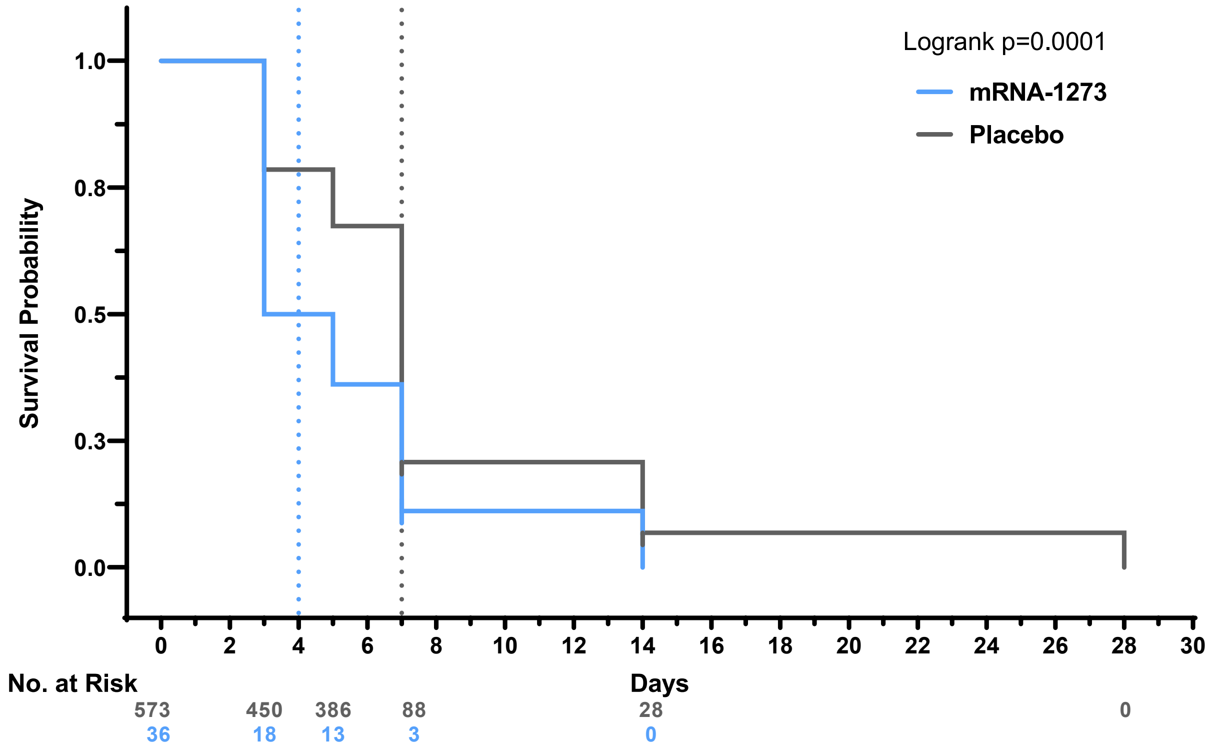

**Dotted lines represent medians (days) for placebo (7) and mRNA-1273 (4). Nasopharyngeal swab at day 1 of illness and saliva samples at days 3, 5, 7, 9, 14, 21, and 28 of illness. Lower limit of quantitation: <2.85 log10 copies/ml for saliva samples**

| Measure | Placebo N=595 | mRNA-1273 N=36 |
|---|---|---|
| Median | 4.0 | 7.0 |
| 1st quartile | 5.0 | 3.0 |
| 3rd quartile | 7.0 | 7.0 |

**Figure S4. Viral copy number CA variants**

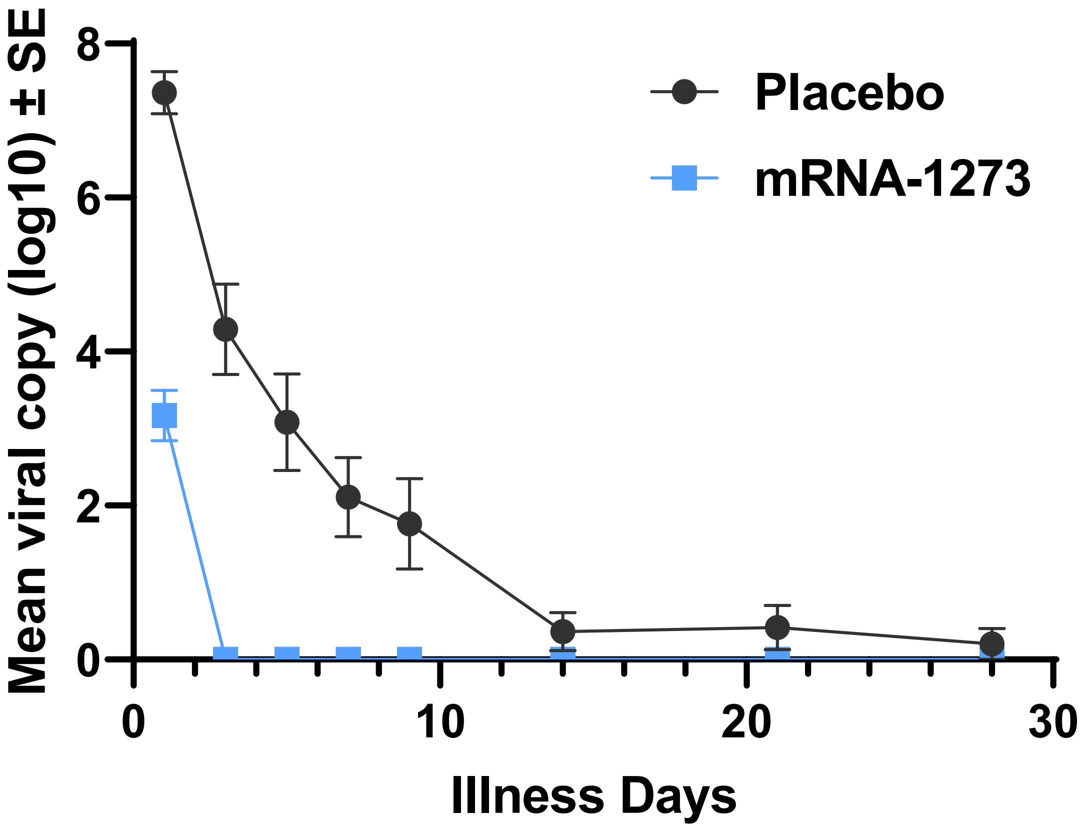

Data underlying graph above.

| Illness Day | mRNA-1273 | | | Placebo | | |
|---|---|---|---|---|---|---|
| | n | Mean | SEM | n | Mean | SEM |
| 1 | 15 | 7.4 | 0.3 | 2 | 6.0 | 1.0 |
| 3 | 14 | 4.3 | 0.6 | 2 | 3.2 | 0.3 |
| 5 | 15 | 3.1 | 0.6 | 2 | 0 | 0 |
| 7 | 15 | 2.1 | 0.5 | 2 | 0 | 0 |
| 9 | 14 | 1.8 | 0.6 | 2 | 0 | 0 |
| 14 | 14 | 0.4 | 0.2 | 2 | 0 | 0 |
| 21 | 13 | 0.4 | 0.3 | 2 | 0 | 0 |
| 28 | 15 | 0.2 | 0.2 | 2 | 0 | 0 |
| SEM=standard error of the mean | | | | | | |

### References

1. Centers for Disease Control and Prevention. SARS-CoV-2 Variant Classifications and Definitions. (Centers for Diease Control and Prevention, 2021).
